# HERITABLE TISSUE-SPECIFIC GENE EXPRESSION ASSOCIATES WITH CHRONIC WOUND MICROBIAL SPECIES

**DOI:** 10.1101/2025.01.22.25320974

**Authors:** Khalid Omeir, Jacob Ancira, Rebecca Gabrilska, Craig Tipton, Clint Miller, Ashley Noe, Kumudu Subasinghe, Megan Rowe, Nicole Phillips, Joseph Wolcott, Caleb D. Philips

**Affiliations:** Department of Biological Sciences, Texas Tech University, Lubbock, Texas, USA; RTL Genomics, MicroGenDX, Lubbock, Texas, USA; Department of Surgery, Texas Tech University Health Sciences Center, Lubbock, Texas, USA; Southwest Regional Wound Care Center, Lubbock, Texas, USA; Microbiology, Immunology & Genetics, University of North Texas Health Science Center, Fort Worth, Texas, USA; Natural Science Research Laboratory, Texas Tech University, Lubbock, Texas, USA

**Keywords:** Chronic wound microbiome, mbTWAS, microbiome-genome association

## Abstract

The reasons for interpatient variability in chronic wound microbiome composition are thought to be complex but are poorly known. To investigate how patients’ genetically regulated tissue expression may influence chronic wound bacterial composition, we performed a microbiome-transcriptome-wide association study. This approach involved estimating for 509 patients their tissue-specific gene expression from DNA genotypes, followed by associating gene expression to the relative abundances of species detected in their wounds as provided on clinical reports to the physician. Comparisons to artery, blood, fibroblast, skeletal muscle, skin, subcutaneous fat, and nerve tissue resulted in 251 transcriptional differences at 109 genes significantly explaining abundances of 39 different species. Overall, these species were detected in ∼63% of wounds. A similar number of associations per tissue was observed (range 31-39), and many genes were associated at multiple tissues in distinct ways. The cumulative variance across loci for species relative abundance explained ranged from ∼3-36%, depending on species. Although the same gene was almost never associated with more than one species, ∼14% of enriched pathways were independently enriched for multiple species, which may reflect the diversity of ways microbes interact with partially overlapping attributes of the wound bed. Commonly enriched pathways pertained to collagen formation and modification, cell signaling, cytoskeletal dynamics, interactions with extracellular matrix, transmembrane proteins, among others. This work expands the new perspective that individual genetics may partially determine microbial colonization and infection.

## 1 INTRODUCTION

Chronic wounds are characterized by failing the normal timescale and progression of healing, with patients commonly suffering wounds for months or years. Chronic wounds are a significant and growing economic burden, but more importantly a humanistic burden that impacts patients’ quality of life. Financially, chronic wounds are responsible for $28-97 billion of Medicare’s annual spending as 15% of Medicare beneficiaries, equivalent to 8.2 million patients, suffer at least one chronic wound.^1^ Patients with chronic wounds commonly exhibit psychoneurological symptoms including cognitive dysfunction, fatigue, depression, and anxiety.^2^ The 5-year mortality rate following ulceration has been reported at 40%.^3^ Overall, patients with chronic wounds are restricted physically and mentally and have a reduced quality of life.

Following wound emergence, microorganisms including bacteria^4^ and fungi^5^ are known to contribute to chronicity.^6^ Reducing bioburden to facilitate healing is a treatment guideline^7^ and is the basis for biofilm-based wound care.^8^ The nutrient-rich wound bed microenvironment provides suitable conditions for microbial attachment and biofilm formation,^9^ and species in biofilms have been shown to be 10-1000 times more resistant to antibiotics in contrast to planktonic states.^4^ Whereas some species are quite common (e.g., *Staphylococcus aureus*, *Staphylococcus epidermidis*, *Pseudomonas aeruginosa*, *Enterococcus faecalis, Finegoldia magna, Anaerococcus vaginalis)*, many species have been observed in chronic wounds and most wounds are polymicrobial.^10, 11^ There is evidence that polymicrobial vs single species infections are more difficult to treat,^12^ which is thought to be a consequence of species’ distinct properties and their interactions.^13^

Microbiomes occurring across human body sites are considered complex traits influenced by the environment and human genetics. Indeed, previous work indicates how genetic differences among people shape microbial communities at different body locations. Linking human genetic variation and microbiome variation is commonly approached by testing the explanatory power of single nucleotide polymorphisms (SNPs) on the variance in abundances of microbes, an approach referred to as microbiome genome-wide association study (mbGWAS). Examples of how mbGWAS have identified loci explaining microbiome composition include the gut,^14^ vagina,^15^ airway,^16^ and chronic wounds.^17^ These discoveries suggest human genetics shapes microbiomes in both health and disease. The prospect that patient genetics explains some of the variation in the chronic wound microbiome suggests avenues to understand patient differences in infection and healing outcomes.

Tipton et al.^17^ demonstrated the potential of mbGWAS to inform potential molecular bases for patient differences in chronic wound microbiomes. Although sample sizes were modest, this work showed through analysis of two cohorts that patients’ genotype at SNP *rs8031916* explained differences in their chronic wound microbiome composition. *rs8031916* is a Talin2 intronic variant, and follow-up comparison of wound bed biopsies showed that patients’ genotype explained differences in the ratio of Talin2 protein-coding alternative transcript expression. These transcripts vary in Talin2 functional domain inclusion. Talin2 is a mandatory mechanosensory protein of focal adhesions, and focal adhesions are important to wound healing^18^ and infection.^19^ Larger studies hold the potential to discover more about how human genetics influences chronic infections. This prospect is encouraging for explaining not only patient differences but may also allow development of individual-level polygenic risk scores for infection (e.g., Pirola et al., 2022).^20^

Because gene expression patterns have high tissue specificity, and multiple tissue types are present in wound beds, framing chronic wound genetic associations in a tissue specific setting could help causally link individual genetics to wound microbiomes. The NIH Genotype-Tissue Expression (GTEx) project^21^ has estimated the heritability of genes in the human genome across a variety of tissues. In doing so, GTEx has provided a mechanism to estimate expression directly from genotype,^22^ which can be used in place of SNPs for association testing. Estimating expression from genotype is valuable in chronic wound research because whereas obtaining DNA from consenting patients is straightforward, accurately dissecting various tissue types from wound beds from the many patients needed for such studies is impractical. To expand our understanding of how patient genetics may influence the microbes present in chronic wounds, a microbiome transcriptome-wide association study (mbTWAS) is presented. For this work, each species was considered an independent phenotype of the patient, thus allowing species-specific analysis and cross-comparison to evaluate emergent trends.

## 2 MATERIALS AND METHODS

### 2.1. Sample Collection

Samples were collected under Western Institutional Review Board protocol 56-RW-156DNA in a deidentified fashion from adults providing written consent for inclusion. Patients with non-healing wounds, persisting for more than three weeks, were recruited during their initial clinical visit to the Southwest Regional Wound Care Center in Lubbock, TX. No exclusion of patients based on chronic wound type (e.g., diabetic ulcers, decubitus ulcers, veinous leg ulcers, arterial ulcers, non-healing surgical wounds) was made. Figure 1 provides a detailed overview of patient exclusion criteria. After initial wound assessment and management planning, wounds were debrided with sharp curette, scissors, and scalpel to remove all slough and devitalized tissue to a bleeding wound bed. The resulting slough and devitalized tissues were manually homogenized with a curette, transferred to a collection tube, and delivered the same day at room temperature to the CAP approved and CLIA certified high complexity diagnostic laboratory, MicroGenDX (Lubbock, TX, USA). In addition to wound tissue debridement, buccal swabs were also collected, which were promptly stored in liquid nitrogen. Buccal swabs were accessioned into the Wolcott Wound Care Research Collection (Museum of Texas Tech University, Lubbock, TX, USA) prior to DNA extraction.

**Figure 1:**
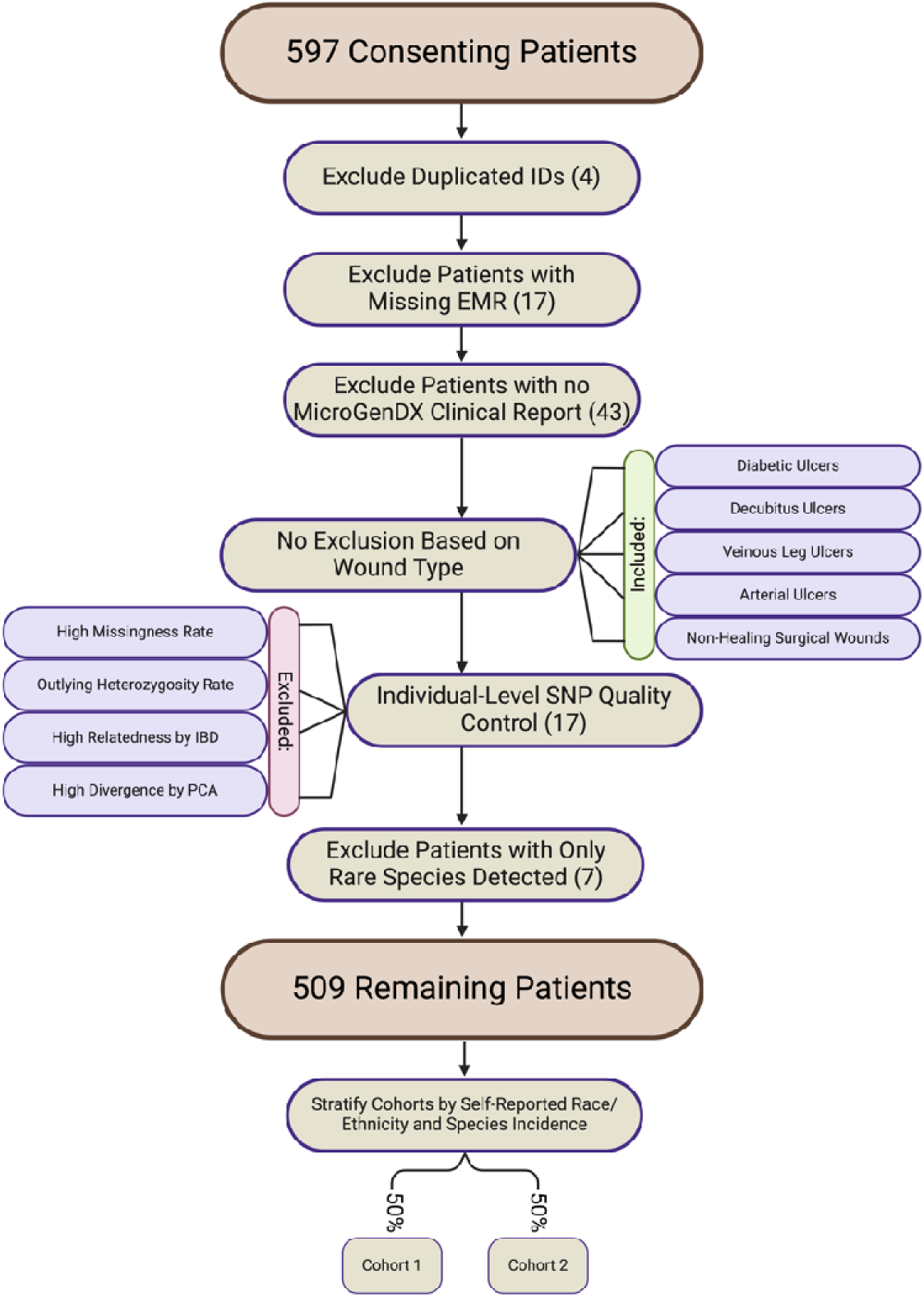
Flowchart of Patient Selection Criteria. There was no exclusion of patients based on patient demographics or wound type. Following exclusion criteria, 88 patients were removed, and 509 patients remained. The number of patients excluded at each step is denoted in parenthetical notation. The cohorts were then stratified by self-reported race/ethnicity and species incidence. Stratification was conducted separately for each species tested. Created using BioRender.com

### 2.2. Patient DNA Extraction and Quantification

DNA extraction from buccal swabs was completed using Gentra Puregene Buccal Kits (Qiagen, Gaithersburg, MD, USA), with overnight lysis at 55°C, inclusion of RNase A for RNA-free purified DNA, and a final elution in 50 μL molecular biology grade water (GE Healthcare Life Sciences MA, USA). DNA samples were quantified using a Qubit 3.0 Fluorometer (Thermo Fisher Scientific, Waltham, MA, USA). All samples were titrated or dehydrated to a final concentration of 10-50 ng/μL. DNA samples were stored at -80°C prior to genotyping.

### 2.3. Patient Genotyping

Genotyping was performed according to the Illumina Infinium High-Throughput Screening (HTS) Assay protocol (Illumina, San Diego, CA, USA). In brief, whole-genome amplification was performed at 37°C for 20-24 hours. Post-amplification, DNAs were fragmented and hybridized to Illumina BeadChips with incubation at 48°C for 16-24 hours. Nonspecifically hybridized and unhybridized DNA were removed by washing, biotin or dinitrophenyl labeled nucleotides were added to extend hybridized primers, and BeadChips (Illumina) were coated for genotype calling. The Illumina iScan System with the GenomeStudio Genotyping Module was used to calculate analysis call rates and heterozygosities. The PLINK Input Report Plug-in extension for GenomeStudio was used to export PLINK files for downstream analyses.

### 2.4. Patient Genomic Data Quality Control

Subjects studied in Tipton et al.,^17^ which were from the same clinic were included. For this, PLINK files were merged based on intersecting SNPs. Strand flipping was performed using --flip flag from Plink 1.9. Multiallelic SNPs and duplicated SNPs were excluded. Illumina GSA support files were used to convert locus names to rsids. Subsequent quality control was consistent with previous protocols^17^ and included omitting patients that exhibited any of (1) high missing data rates (> 0.09), (2) outlying heterozygosity (±3 standard deviations from mean), (3) high levels of relatedness evident by an identity by descent of more than 0.1875, or (4) highly divergent ancestry exhibited through principal component analysis using the SNPRelate package.^23^ SNP/Marker-level filtering was performed to omit any loci with (1) low genotyping rates (<0.05), (2) any SNP out of Hardy-Weinberg equilibrium (p < 1E-5), or (3) a minor allele frequency <0.05.

### 2.5. Genome and Transcriptome Imputation

Following quality control, genome imputation was performed to increase loci inclusion. Eagle 2.4.1^24^ was used to phase the genotypic data followed by Beagle 5.4^25^ to impute missing genotypes. The haplotypes were estimated using the corresponding hg37 reference map. The 1000 Genomes Project phase 3 reference panel was used to impute the genome. Imputed SNPs with a dosage *R*^2^ less than 0.80 were omitted. SNPs were queried using USCS’s bigBedNamedItems tool to convert to hg38 build.^26^ To account for imputation uncertainty, the dosage values of SNPs were used to impute the genetically regulated expression via PrediXcan^27^ in Python release 3.5.0. Expression and splicing MASHR-based prediction models, which are parsimonious and biologically informed, trained on the GTEx v8 release were downloaded from predictDB.^27, 28^ PrediXcan (Predict.py) was used to impute the gene-level expression and intron-retention splicing patterns for a priori defined biologically relevant tissue types including artery, blood, fibroblast, skeletal muscle, skin, subcutaneous fat, and nerve tissue. The sampling sites of tissues performed by the GTEx Consortium were as follows: 1) peripheral tibial artery, 2) femoral vein/subclavian vein/heart, 3) cultured primary fibroblasts, 4) gastrocnemius muscle, 5) left or right leg 2 cm below patella on medial side, 6) subcutaneous tissue beneath the leg’s skin, and 7) peripheral tibial nerve, respectively.^21^ Mapping rules were specified using --on_the_fly_mapping option because MASHR models include many variants that lack rsids.

### 2.6. Chronic Wound Microbial Compositional Data

Chronic wound bacterial species relative abundance profiles were retrieved from the clinical reports provided to the physician by MicroGenDX. In brief, the clinical reports were generated as follows: DNA from wound samples was extracted using a Kingfisher Flex automated platform (Thermo Fisher Scientific, Waltham, MA, USA) and ZymoBIOMICS-96 MagBead DNA extraction reagents (Zymo, Irvine, CA, USA). Amplification of bacterial 16S V1-V2 was performed using ABI Veriti thermocyclers (Applied Biosystems, Carlsbad, CA, USA) using primers 28F (GAGTTTGATCNTGGCTCAG) and 388R (GCTGCCTCCCGTAGGAGT). PCR products were pooled based on agarose gel banding intensity and pools were size selected using two rounds using Agentcourt AMPure XP (Beckman Coulter, Indianapolis, IN, USA). The pools were quantified using a Qubit 2.0 Fluorometer (Thermo Fisher Scientific, Waltham, MA, USA) and pooled libraries were sequenced on Illumina MiSeq. Following quality control and assembly, reads were clustered into operational taxonomic units (OTUs) at a 97% sequence similarity. OTU assignment was performed using an in-house curated taxonomic reference database. Extractions and PCR controls were implemented to validate results and detect contamination by clinical lab supervisors. Only species with a relative abundance greater than 2% were included on the clinical reports. Those molecular methods have recently been described in detail elsewhere.^17, 29^ The study-wide incidence rate of each species was tabulated, and any with an incidence rate of at least 1% was retained. Prior to association testing, the relative abundance distribution of each species across patients was centered-log ratio (CLR) transformed. Subsequent association testing was conducted separately for each species, with their CLR transformed relative abundances considered the phenotypic measure.

### 2.7. Association Testing

The predicted genetically regulated expression of genes within each tissue was linearly associated to CLR transformed species abundances using PrediXcan (PrediXcanAssociation.py) including age, diabetes, sex, eigenvector 1 (EV1), and eigenvector 2 (EV2) as covariates (Figure 2). Inclusion of EV1 and EV2 to control for genetic ancestry was determined by first conducting variance normalized principal component analysis in plink with option --pca. The explanatory power of eigenvectors on racial category was then assessed using PERMANOVA with Euclidean distances in which the explanatory power of vectors 1-10 on a 0/1 coded self-identified race/ethnicity matrix was assessed. EV1 and EV2 were significant and explained ∼86% of variance, whereas eigenvector 3 was non-significant and eigenvectors 3-10 cumulatively explained only 0.2% of remaining variation. Therefore, EV1 and EV2 were included as covariates in subsequent association testing, which is consistent with expectations of K -1 (K = number of populations) significant vectors demonstrated by Patterson et al.^30^

**Figure 2:**
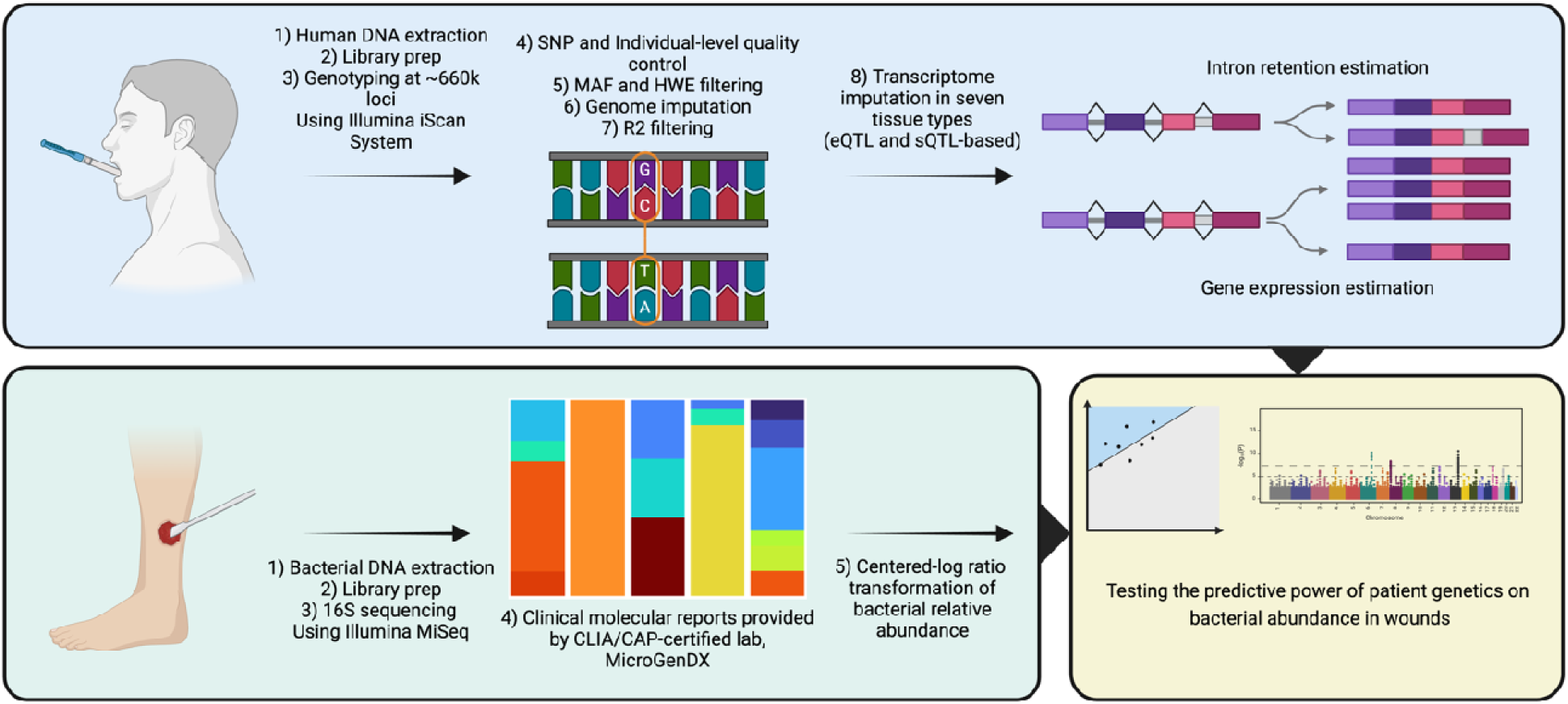
Summary of Experimental Design. Top panel: DNA extracted from buccal swabs was used to genotype patients, from which gene and intron retention levels were estimated from GTEx models. Bottom left panel: Species relative abundances were measured from homogenized wound debridement by the clinical lab. Bottom right panel: The explanatory power of tissue specific gene expression to explain species’ relative abundances was tested with linear modeling. Created using BioRender.com

For each phenotype, associations were performed using two cohorts with balanced sample sizes and stratification on self-identified race/ethnicity and species incidence rates. Bonferroni multiple testing corrections were used to identify candidate significant associations in each cohort, then for that limited subset of loci their repeated significance in the other cohort was evaluated with multiple testing correction.

For species with more than one significant association the additive effects across loci were assessed using multiple regressions including all loci and covariates with backward model selection based on p-value. The adjusted *R*^2^ for each model was recorded, and the relationship between *R*^2^ and number of associated genes across species was evaluated with regression. For genes associated to the same species across different tissues, the distinctiveness of expression for a given gene in each tissue was determined based on if multiple predictor terms corresponding to different tissues were retained in the pruned model.

Binomial tests were used to assess if genera were significantly overrepresented among the set of associated species in reference to the full list of species tested. Genera with at least one associated species were included for evaluation. The number of trials was set to the number of species for a given genus, the number of successes was set to the number of species with significant associations for a given genus, and the a priori hypothesized probability of success was the frequency of a given genus in the overall list of species included for association testing. A Bonferroni multiple testing correction was employed across tests prior to interpretation.

### 2.8. Functional Enrichment Analysis

Overrepresentation analysis of Reactome pathways^31^ was conducted using R package clusterprofiler.^32^ Given the number of species, tissues, classification of species based on aerotolerance, and delineation of associations at the gene or transcript level, there were many possible ways enrichment analyses could be focused. Here, analysis was conducted separately on associated gene lists grouped as a) study-wide associated genes, b) genes grouped by tissue irrespective of species, c) genes grouped by species irrespective of tissues, d) genes grouped by individual tissue-species pairs, and e) genes grouped by aerotolerance-tissue combinations (for the latter, genes associated to individual species were combined into their respective aerotolerance classification of aerobic, facultative anaerobic, and anaerobic). Significant enrichment was defined using Benjamini–Hochberg multiple testing correction maintaining α < 0.05. To meet clusterProfiler and reactomePA input requirements, ENSG IDs were converted to ENTREZ IDs using the function bitr(). The background gene sets for overrepresentation analysis were comprised of all successfully imputed genes for tissues relevant to each comparison.

## 3 RESULTS

### 3.1. Patient Demographics and Wound Profiles

After quality control, the study sample consisted of 509 patients (309 males, 200 females) retained for association testing. The average age of participants was 68 years and ∼19% identified as smokers. The majority of patients had diabetes (325 patients; ∼64%) and were mostly overweight or obese (420 patients; ∼83%). The self-described ethnic/racial distribution was 38 “Black”, 142 “Hispanic”, and 329 “White”. The most common demographic of the study was self-described white, obese, non-smoker males. Sixty-eight bacterial species were observed in at least 1% of patients.

### 3.2. Quality Control, Genomic and Transcriptomic Imputation

After SNP level quality control from the genotyping array, 306,942 SNPs remained with a genotyping call rate of 0.996971. Genotype imputation provided an initial set of 30,761,485 SNPs, which were then filtered to high confidence loci based on *R^2^* resulting in 12,094,556 SNPs retained. Based on these SNPs, tissue-specific transcriptome imputation predicted an average of 12,065 genes and 30,721 splicing events per tissue (Table 1).

**Table 1:** Number of successfully imputed genes and intron retention events per tissue.

| Tissue | Genes | Intron Retentions |
| --- | --- | --- |
| Artery | 12,218 | 33,274 |
| Blood | 10,591 | 19,669 |
| Fibroblast | 11,923 | 29,866 |
| Skeletal muscle | 11,283 | 25,708 |
| Skin | 12,896 | 34,365 |
| Subcutaneous fat | 12,407 | 34,928 |
| Nerve Tissue | 13,137 | 37,237 |

### 3.3. Association Testing

Out of 476 independent species-tissue association studies 157 (∼33%) yielded at least one association, which was defined as a locus that was significant in both cohorts with multiple testing correction thresholds. There were 251 associations at 109 genes, with ∼66% of these being to intron retention (166 associations; 65 unique genes), and ∼34% being to gene level expression (85 associations; 45 unique genes) (Figure 3). There was no relationship between species incidence rate and number of significant gene associations (p = 0.1). No genes were associated with the four most commonly detected species *Staphylococcus aureus*, *Staphylococcus epidermidis*, *Finegoldia magna*, and *Escherichia coli*, but 39 other species had at least one significant association. This set of 39 species had a median incidence rate of 3%, and collectively, they were detected in the majority of patients’ wounds (323, ∼63%). With respect to the full list of tested species, the genera *Staphylococcus* (p = 0.002), *Anaerococcus* (p = 0.02), *Prevotella* (p = 0.02), and *Klebsiella* (p = 0.02) were overrepresented among the list of associated species.

**Figure 3:**
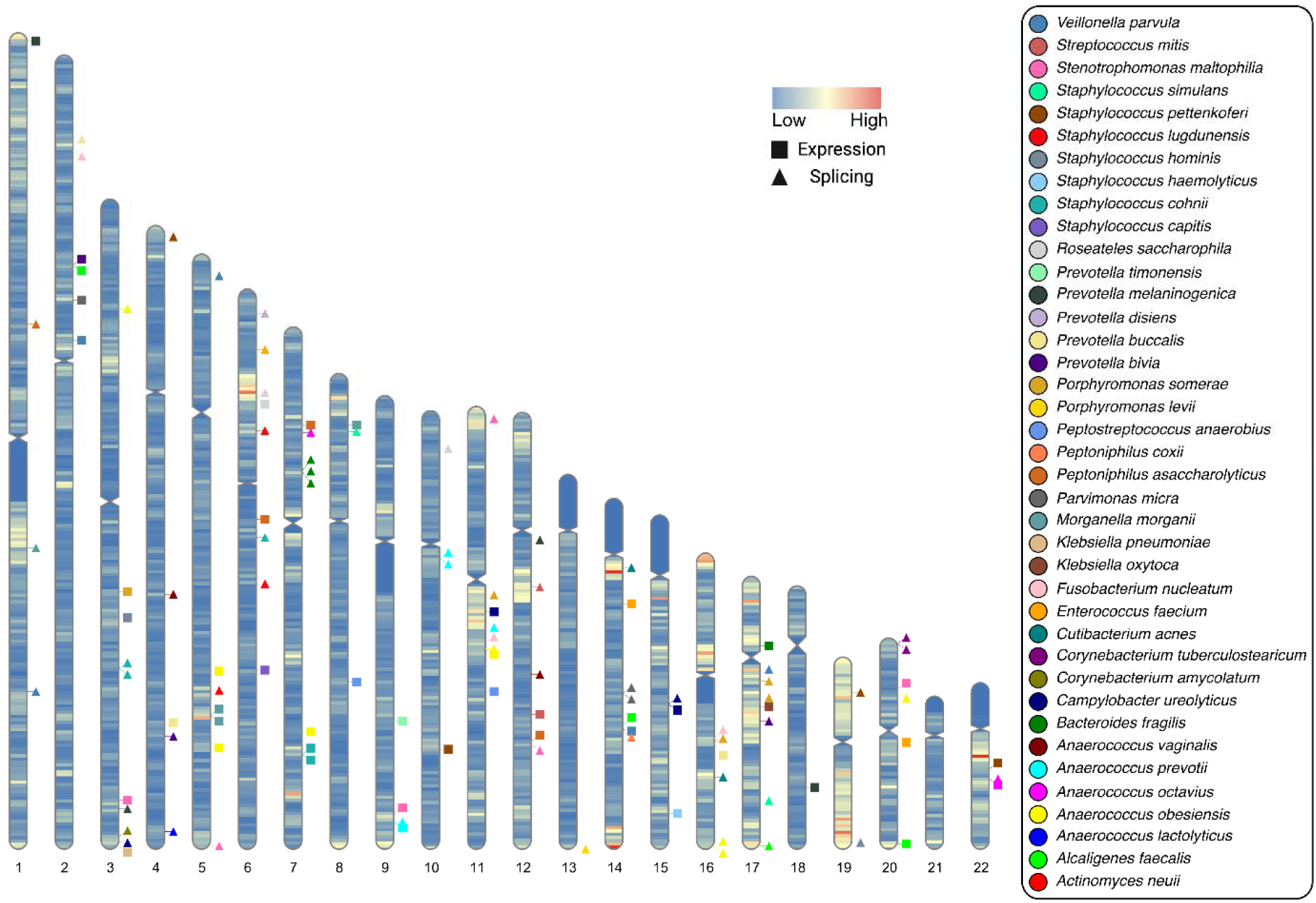
Chromosomal ideogram illustrating the 109 significant genes identified through two cohort mbTWAS analysis. Each marker represents the mid-position of a significant gene on chromosomes directly to the left. Marker colors indicate the species to which genes were associated, and shapes indicate if the association was for a gene or intron retention. Chromosomal gene density is illustrated by banding. (R: RIdeogram^73^)

The median and modal number of associations was five (Q1 = 3, Q3 = 8.5) and three, respectively (Figure 4), and 28 (∼72%) of the significant species were associated to two or more genes. *Anaerococcus obesiensis* had the largest number of associations (34), which were distributed across eight genes. To evaluate if significant associations had additive effects for explaining species relative abundances, covariates and significant loci were jointly considered in multiple regression with backwards selection. Through these analyses explanatory power on species’ abundances ranged between 3-38%, and the explanatory power of models for each species was strongly influenced by the number of loci associated to species (Figure 5).

**Figure 4:**
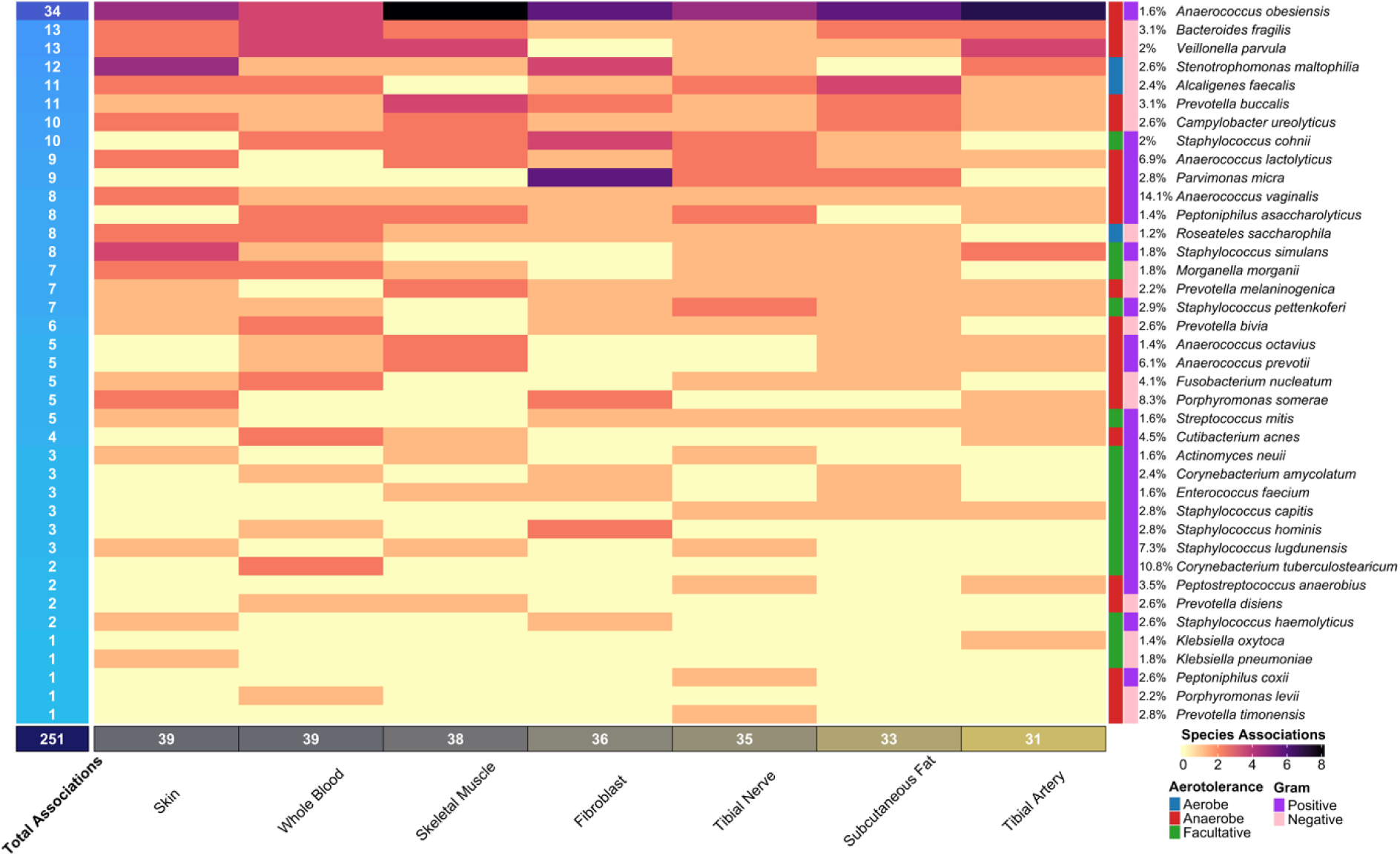
Heatmap illustrating the number of significant associations by tissue and species, with tissues as columns and species as rows. Leftmost column reports total associations for each species and is shaded proportional. Righthand columns illustrate from left to right aerotolerance, Gram staining, and incidence rate. The bottom row reports column totals. (R: ComplexHeatmap^74^)

**Figure 5:**
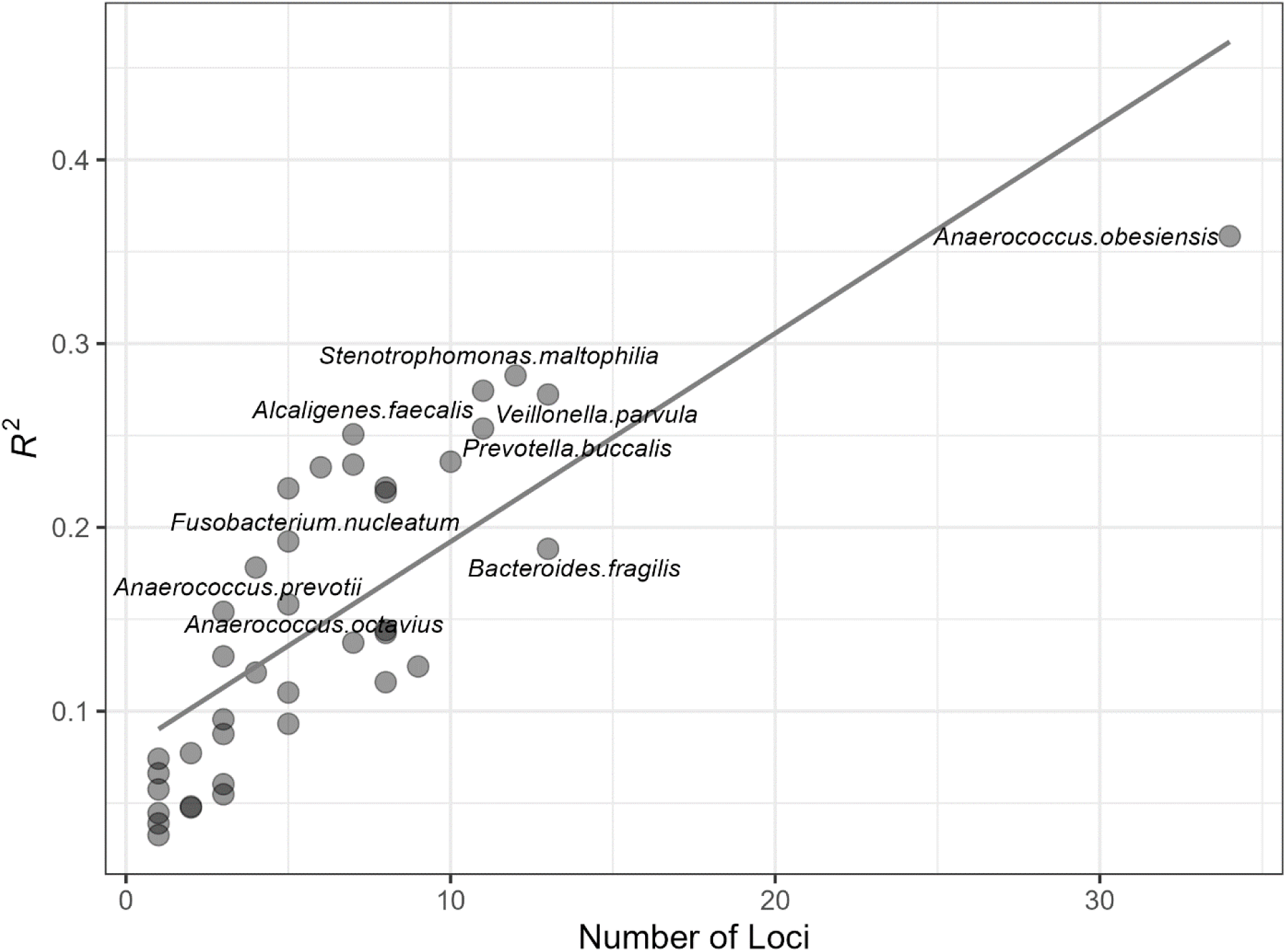
Regression plot illustrating the relationship between adjusted R^2^ and the number of loci that were significantly associated to each species (F = 58, df = 1,37, p < 0.001). Points denote individual species, and some points are not labeled to reduce clutter.

The number of associations per tissue ranged from 31 to 39. Only one gene (*VPS54*; VPS54 subunit of GARP complex) was found to be significantly associated with different species (*Alcaligenes faecalis* and *Prevotella bivia*). Reflecting how some SNPs predict a gene’s expression in multiple tissues, albeit potentially in different ways, 191 of the 251 (∼76%) associations were replicated in multiple tissues. These multi-tissue associations were distributed across 53 unique genes. Multiple regression model selection indicated that ∼42% of these genes (22) were associated to their corresponding species in significantly distinct ways across tissues. The most common differentially expressed gene was *SYNPO* (synaptopodin), observed six times in association to *A. obesiensis* (all tissues except artery; Figure 7). *ACSL1* (acyl-CoA synthetase long chain family member 1) had the most differential intron retention associations with nine to *A. lactolyticus*. The intron at chr4:112432581-112435148 of *ALPK1* (alpha kinase 1) was the single intron with the most associations, and its decreased excision was associated with increased abundance of *A. vaginalis* across tissues (see Supplementary Table 1 for full reporting of significant associations).

**Figure 6:**
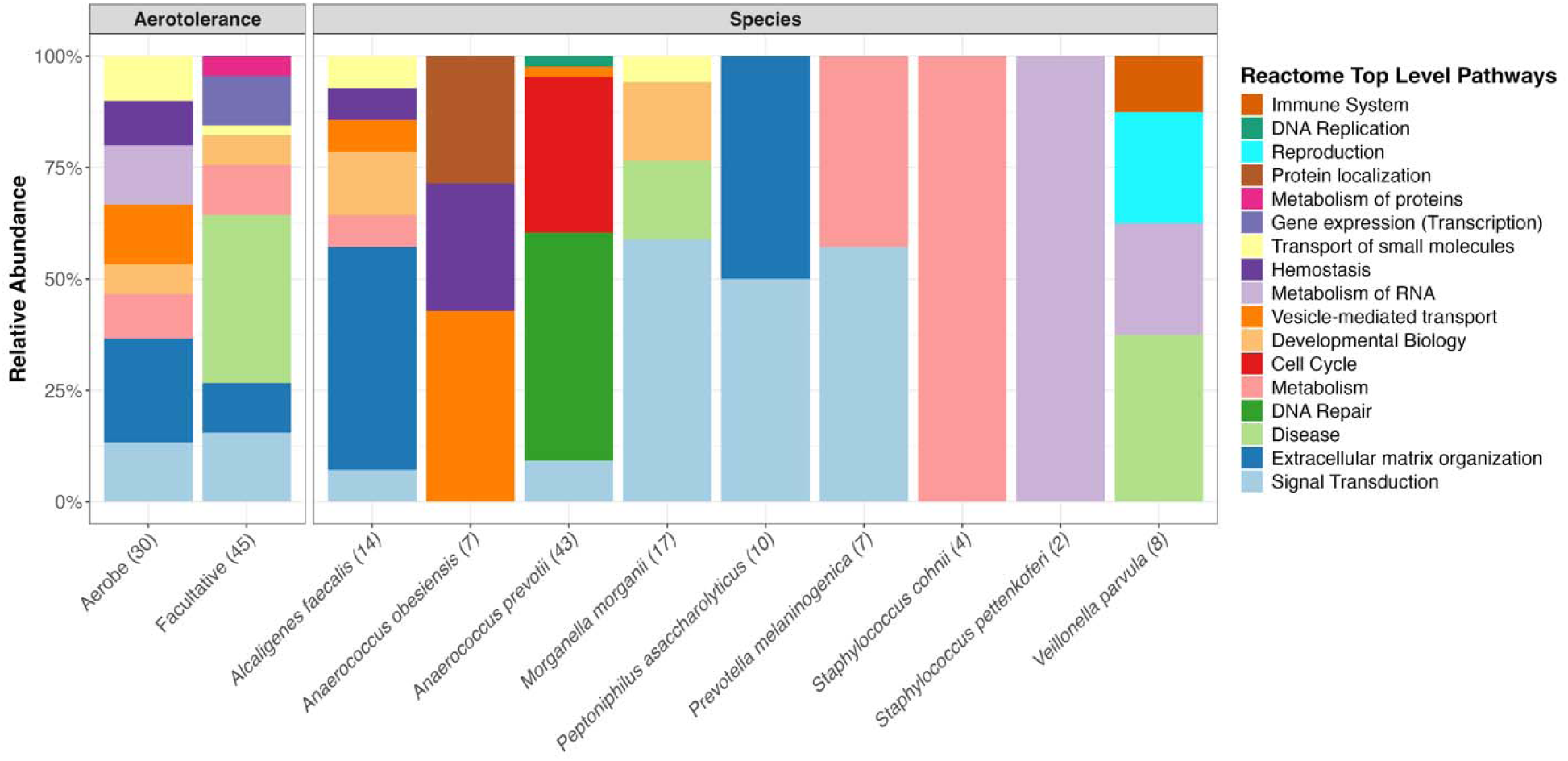
Pathway enrichment results for aerotolerance (left) and species (right) summarized as relative abundances of Reactome Top Level Pathways.

**Figure 7:**
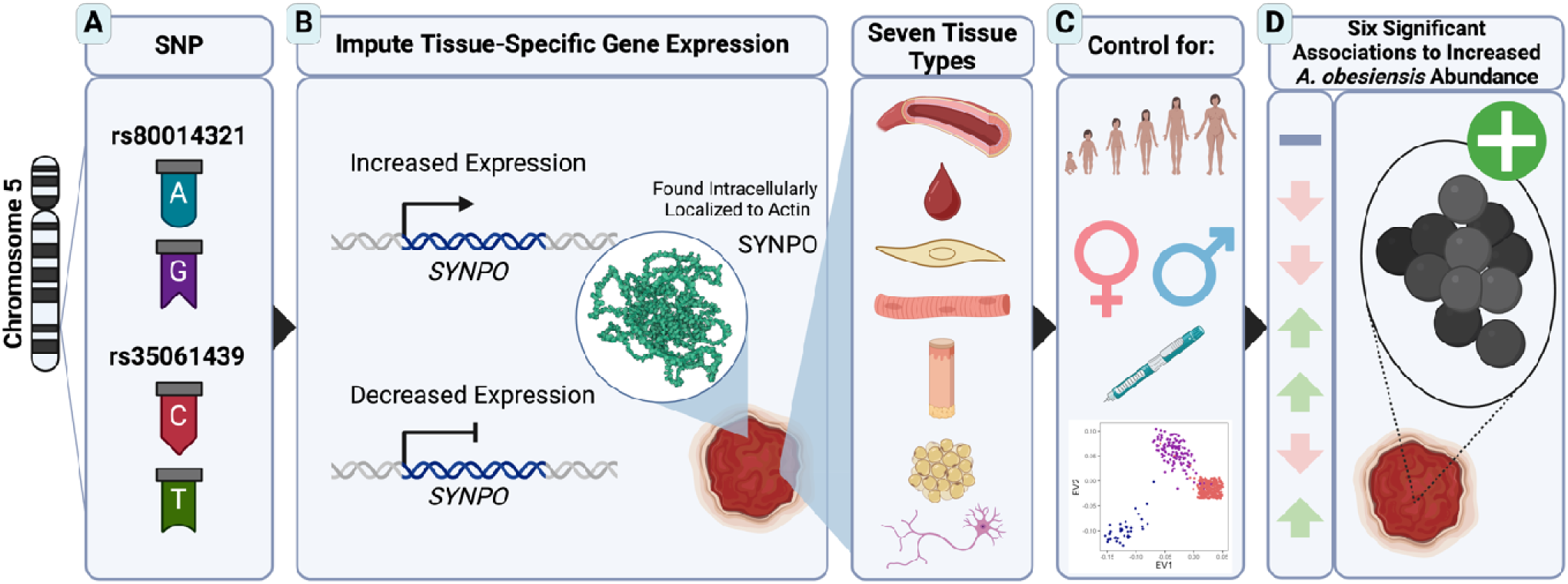
Schematic Example Illustrating Relationship Between Patient Genetics and Ultimately Microbial Species. (A) Two SNPs on chromosome 5 (rs80014321 and rs35061439) are predicted to influence tissue-specific expression of the SYNPO gene. (B) The genetically regulated expression in seven different tissues (artery, blood, fibroblast, skeletal muscle, skin, subcutaneous fat, and nerve tissue; illustrated top to bottom) was imputed for patients based on their genotype. (C) Association testing controlled for: age, sex, diabetic status, and genotypic eigenvectors (illustrated from top to bottom). (D) Variation in genotypically predicted SYNPO expression was significantly associated with observed relative abundance of A. obesiensis across six of seven tissues. Increased or decreased SYNPO expression in association to A. obesiensis abundance is presented by upward green or downward red arrows, and non-significance by a minus sign. SYNPO structure obtained from AlphaFold.^75^ Created using BioRender.com

### 3.4. Enrichment Analysis

Functional enrichment analyses for different groups of associated genes were next performed, and Supplementary Tables 2-4 provides a full reporting of these results. First, all study-wide associated genes were simultaneously tested through which no significantly enriched pathways were identified. Testing gene lists grouped corresponding to tissue irrespective of species also resulted in no enrichments. However, gene lists grouped by species irrespective of tissue did yield significance. Specifically, among 13 species with enough associations to allow analysis, nine were associated to enriched pathways encompassing 113 significant terms. These species and their corresponding number of enriched terms were *A. prevotii* (43), *M. morganii* (17), *A. faecalis* (15), *P. asaccharolyticus* (10), *V. parvula* (8), *P. melaninogenica* (7), *A. obesiensis* (7), *S. cohnii* (4), and *S. pettenkoferi* (2). Summarizing to Top Level Reactome Pathways, “signal transduction”, “extracellular matrix”, and “disease” were most common, but each species had a unique set of associated terms (Figure 6). Fourteen percent of pathways were independently significant for multiple species. Overlapping pathways could be further summarized to a) tRNA (“tRNA modification in the nucleus and cytosol”, “tRNA processing”), b) collagen (“Collagen chain trimerization”, “Collagen degradation”, “Collagen biosynthesis and modifying enzymes”, and “Collagen formation”), and c) RHO GTPase cycle (“CDC42”, “RHOJ”, “RHOQ”, “RHOB”, “RHOC”, “RHOA”, and “RAC1 GTPase cycle”). CDC42 GTPase cycle was coenriched most often (three; *A. prevotii*, *M. morganii*, *P. melaninogenica*; Supplementary Table 2).

Performing the same analysis but narrowed to individual tissue-species pairs yielded 26 pairs with significantly enriched terms, albeit only two species, *A. obesiensis* and *S. maltophilia*, were involved. *A. obesiensis* accounted for the majority (∼77%) of these enrichments that were distributed across five tissues, and mitochondrial protein import and protein localization were repeatedly enriched across tissues (Supplementary Table 3).

Enrichment based on aerotolerance-tissue pairs yielded no functional enrichment for anaerobe associated genes. However, significant enrichments for both facultative anaerobes and aerobes were detected. For facultative anaerobes (represented by 15 species, Figure 4), there were 45 enriched terms distributed across artery (25), skin (15), and fibroblast (5) tissues. For aerobes (represented by *Alcaligenes faecalis, Roseateles saccharophila*, and *Stenotrophomonas maltophilia*) there were 32 enriched terms distributed across adipose (15), nerve (9), artery (4), skeletal muscle (3), and whole blood (1) tissues. There was minimal tissue overlap between facultative and aerobic enrichments, only occurring at arterial tissue. However, overall, they were coenriched for collagen-related terms (collagen chain trimerization, collagen degradation, assembly of collagen fibrils and other multimeric structures, collagen biosynthesis and modifying enzymes). Collagen-related terms were enriched in subcutaneous fat for aerobes and in artery for facultative anaerobes. Also, each group was enriched for different terms pertaining to cell surfaces with facultative anaerobes enriched for spike protein, phagocytosis, and non-integrin extracellular interaction terms, and aerobes enriched for extracellular matrix and integrin-related terms, among others (Supplementary Table 4). About half (5 of 11) Top Level Reactome Pathways were shared (Figure 6).

## 4 DISCUSSION

This work was motivated by the general hypothesis that clinically relevant differences in the microbes observed among chronic wounds may be determined in part by patient genetics. Support for this hypothesis is multifaceted. Generally, that individuals can have genetic predispositions to infectious disease has been known for quite some time.^33^ Also, there are multiple examples of how peoples’ genetic variation associates with microbiome variation across many body sites including skin,^34^ and wounds are thought to be typically colonized from adjacent healthy skin.^6^ A relevant example is the evidence for a genetic predisposition to an opportunistic pathogen provided by Brown et al.^35^ who reported that categorizing people as non-, intermittent-, or persistent-carriers of *S. aureus* in healthy nasal passages was genetically associated.

Tipton et al.^17^ provided the initial results supporting a patient genetic effect on chronic wound microbiome characteristics. That work treated a measure of wound microbiome diversity (Hill1, i.e., the exponentiation of the Shannon Weaver Diversity Index) as the phenotype associated to human genetics. Treating diversity as the associated phenotype was innovative in that it greatly simplified comparisons and provided a continuous variable summarizing microbiome. However, it is not immediately apparent how cell characteristics can directly select for microbiome diversity. Post hoc investigation suggested that differences in prevalence of key species (*Pseudomonas aeruginosa* and *Staphylococcus epidermidis*) may underly the genetic association to diversity via their roles in influencing community composition. Herein, to directly evaluate the association of human genetic variation with species, each species was considered a distinct phenotype. Although how associated species contributed to the bioburden in the wounds studied here is unknown, most of these species have been reported as opportunistic pathogens. A non-exhaustive list of such species includes *Alcaligenes faecalis*,^36^ *Anaerococcus lactolyticus*,^37^ *A. octavius*,^37^ *A. prevotii*,^37^ *A. vaginalis*,^37^ *Bacteroides fragilis*,^38^ *Campylobacter ureolyticus*,^39^ *Cutibacterium acnes*,^40^ *Fusobacterium nucleatum*,^41^ *Klebsiella oxytoca*,^42^ *Morganella morganii*,^43^ *Parvimonas micra*,^37^ *Peptoniphilus asaccharolyticus*,^37^ *P. coxii*,^37^ *Peptostreptococcus anaerobius*,^37^ *Prevotella melaninogenica*,^44^ *Staphylococcus capitis*,^45^ *S. cohnii*,^46^ *S. pettenkoferi*,^47^ *Stenotrophomonas maltophilia*,^48^ and *Veillonella parvula*.^49^

Of the 109 associated genes, 108 were associated with single species. This broad independence of associated genes suggests diversity in how species interact with the wound bed. Investigating gene functions for individual species associations often provided clues about the basis for the association. For example, the gene with the most associations to *A. obesiensis* was the actin-binding protein SYNPO. Actin structures can be exploited during cellular invasion^50^ and SYNPO is thought to be important for wound healing.^51^ Moreover, bacteria can mediate cellular invasion through actin-mediated phagocytosis.^52^ Additionally, bacteria can adhere to the host cell via microtubule wrapping.^52^ *KLC4* (kinesin light chain 4), a microtubule-associated force-producing protein,^53^ was found associated to *Actinomyces neuii* in skeletal muscle. *A. vaginalis* had the most associations to *ALPK1*, which is involved in detection of bacterial metabolites, plays roles in cytoskeleton and non-motile primary cilia,^54, 55^ and was previously reported to be activated by an anaerobe.^56^ Upon detection of bacterial metabolite, ALPK1 triggers a signaling cascade, resulting in production of proinflammatory cytokines and chemokines.^56^ Another associated cytoskeletal gene was *INA* (internexin neuronal intermediate filament protein alpha), for which its decreased expression in fibroblast and nerve tissue was associated with increased abundance of *S. pettenkoferi*. Epithelial intermediate filaments form a structural barrier against microbial infection and can trigger inflammatory responses.^57^ While no evidence of neuron-specific intermediate filaments interacting with microbiome exists to our knowledge, they contain a general structure shared by all intermediate filament proteins.^58^ The only gene associated with more than one species (*A. faecalis* and *P. bivia*) was *VPS54* that is involved in Golgi-associated retrograde protein transport, which is a mechanism of bacterial toxin entrance,^59^ which may be exploited by these species in yet to be characterized ways.

Above examples provide a non-exhaustive summary of known roles of some associated genes and suggest aspects of how each species may uniquely interact with human tissue. However, by comparing pathway enrichments across species, commonalities also emerged. Among shared pathways (i.e., those that were enriched for more than one species) collagen formation and RHO GTPase cycle related terms dominated. Collagen is the most abundant protein in ECM. ECM, collagen, and integrin pathways are critical in wound healing^60^ and play a role in infection by affecting bacterial establishment and spread.^61^ Also, the RHO family GTPases are essential for various cellular processes that include actin organization, cytoskeleton reorganization, cell adhesion and migration, gene expression, mitosis, morphogenesis, polarity, and vesicle trafficking.^62^ Previous work elucidates how bacteria exploit GTPase signaling for entry and survival.^63^ Also, the CDC42 GTPase cycle, which was enriched across the most species, is a known regulator of cytoskeletal and epithelial morphogenesis by coordination of adhesion.^64^ Mechanistically, bacteria can bind to integrins using their fimbriae or other adhesins, leading to the formation of focal adhesion complexes and activation of RHO and CDC42, along with other proteins.^50^

Summarizing enrichments by grouping species into aerotolerances additionally emphasized pathways related to cell surface interactions. Collagen-related terms were the only ones to be enriched for multiple aerotolerances, which was based on associations of collagen alpha chains to *A. faecalis* and *S. capitis*. Interestingly, associations to different alpha chain genes conferred enrichment of both integrin (collagen type IX alpha 3 chain) and non-integrin (collagen type X alpha 1 chain) cell surface interactions. So, although there was diversity in the types of genes associated, prevalence to aspects of the cytoskeletal-cell surface-ECM architecture is expected given they are primary interaction points with bacteria. This pattern is also consistent with Tipton et al.^17^ who hypothesized that genetically regulated differences at focal adhesions are consequential to wound microbiome differences. Focal adhesions may also be stabilized by *FNBP1* (formin binding protein 1), which can also regulate cell migration^65^, and an intron retention in *FNBP1* was found associated to increased *A. prevotii* abundance in arterial tissue.

Whereas grouping genes by species irrespective of tissue resulted in several enriched pathways, grouping genes by tissue irrespective of species yielded no enrichments. This indicates a lack of tissue-specific attributes that differentially select for multiple species. Related, ∼76% of associations occurred in more than one tissue; that is, species were commonly associated to the expression of a given gene across multiple tissues. Such repeated associations could be caused by a lack of tissue-specificity of a gene’s expression resulting in the exact same association test being repeated across comparisons. Alternatively, the gene’s expression could vary across tissues, and each tissue’s gene expression distribution being uniquely associated to the species. Results of model selection for loci predicting species’ expression indicate that both possibilities occur. Among genes that had multiple associations to a species, ∼42% were included in the final models as multiple significant predictors reflecting their unique effects within each tissue.

One trend that stands out is that no loci were significantly associated to the most common species observed in this study (*Staphylococcus aureus, Staphylococcus epidermidis*, *Finegoldia magna*, and *Escherichia coli*). This could be due to chance cohort structure, their relative abundances being too variable to obtain significant associations, or due to these species acting as generalists with respect to substrate selection. The latter possibility seems plausible given their high incidence rate. However, Brown et al.^35^ reported a genetic association to *S. aureus* in healthy subjects, and Tipton et al.^17^ indirectly suggested an association to *S. epidermidis* in wounds. It is also possible that any patient genetic selection for these species is distributed across many loci of modest additive effects, which would be difficult to detect with the large multiple testing corrections used herein. In any case, given the importance of these species in wounds follow-up investigation is warranted, for example by conducting association analysis similar to that reported herein, but focusing on species prevalence rather than relative abundances.

Another notable pattern from the presented results was that certain genera had more associated species than expected by chance. For example, seven of 10 *Staphylococcus* species (p = 0.002) and four of six *Anaerococcus* species (p = 0.02) tested had significant associations. Disproportionate representation of a genus among associated species may indicate phenotypic properties related to colonization capability that are shared among congeners and selected by wound substrate. However, no congeners were associated with the same gene. For the reduced set of species for which enrichment analysis was possible, only the term “vesicle-mediated transport” was shared between *A. obesiensis* and *A. prevotii*. Another possibility is that substrate variability is important to the colonization potential of such genera and their species tend to diversify for substrate preference. In any case, re-evaluating this trend in larger and independent cohorts would be needed to further evaluate this phenomenon.

Several of the genes identified in this study have also been implicated in other host-microbiome contexts. Using the GWAS catalog^66^ (accessed September 22, 2024), seventeen of the significant genes in the current study (Supplementary Table 5), including *ALPK1*, *KIF16B* (kinesin family member 16B), *MMP15* (matrix metallopeptidase 15), *POLM* (DNA polymerase mu), *SENP6* (SUMO specific peptidase 6), *TUSC3* (tumor suppressor candidate 3), and *VPS54*, were reported as linked to significant SNPs from other mbGWAS. The overlap in associated genes among studies may highlight a shared genetic basis involved in microbial interactions across body sites other than wounds, suggesting that genetic determinants may influence host-microbiome interactions in a broader, systemic context. Two example genes include *KIF16B* and *SENP6*, which were not only significant herein, but were also mapped by SNPs significant to both the gut^67, 68^ and vaginal^69^ microbiome. *KIF16B* regulates *MT1-MMP*, a matrix metalloproteinase with roles in leukocyte extravasation and inflammation, as well as ECM degradation in macrophages.^70^ *SENP6* regulates toll-like receptor signaling, which is critical for host immunity and response to microbial invasion.^71^ Collectively, results suggest the influence of patient genetics on wound microbiomes is simultaneously wound-specific and distributed across body sites.

In conclusion, microbes in wounds are known to exacerbate chronicity, and this work greatly expands our emerging understanding of the role patient genetics may play in shaping wound microbiomes. Given species differ in their pathogenicity and recalcitrance to treatment, such genetic effects may be consequential to health outcomes. Also, the functional plausibility of genes and enriched pathways provided here can be useful to form hypotheses about species’ mechanisms of interaction. Moreover, seeing that the inclusion of additional loci provided a linear additive effect on explaining species’ abundances suggests that there may be many undiscovered causal loci that can be revealed with additional and larger cohorts. It is also noteworthy that despite potential effects from multiple uncontrolled variables, repeatable associations were discovered through the two-cohort design. For example, samples were collected at single timepoints, and previous work has demonstrated wound microbiomes can be compositionally variable over time.^72^ Also, patients’ wounds certainly had many unknown historical differences in treatment prior to attending the participating wound clinic for the first time. The slough and devitalized tissue from which microbiome profiling was conducted is expected to contain some number of dead bacteria and extracellular DNA that could introduce additional microbiome compositional variance, still many associations were repeated across cohorts. These observations indicate that the relationship between patient genetics and wound microbiome is reliable, and future longitudinal models incorporating microbiome measured at multiple timepoints and locations in the wound bed can further enhance understanding how patient genetics determines wound microbiome composition.

Understanding in detail how patient genetics influences microbes in wounds not only lends mechanistic insights that themselves may inform novel treatment approaches but also may provide the opportunity to develop individualized polygenic risk scores. These scores, which would indicate a person’s genetic predisposition to infection by specific species, could have utility both prophylactically and interventive. For example, risk scores could inform prophylactic antibiotic administration prior to surgery. Such scores could also inform why some chronic wound sufferers have consistent bioburden for specific species and indicate the need for personalized intervention including NGS-guided topicals. It is also foreseeable that in the future patient genetics may associate not only with specific species, but also with healing outcomes. The ability to predict a patient’s likely healing trajectory under standard-of-care could serve as a reference point to gauge the effectiveness of personalized intervention.

## Data Availability

The genomic data used for this study is available in the Gene Expression Omnibus (GEO) repository with the accession number: GSE276944.

## ACKNOWLEDGEMENTS

We thank MicroGenDX and CEO Rick Martin for their support of this work. We thank the staff at the Southwest Regional Wound Care Center for their commitment to excellence in wound care, scientific collaboration, and logistical support. This work was supported by NIH award R15GM141973-01.

The authors Khalid Omeir, Jacob Ancira and Craig Tipton are employed by, and Caleb Phillips is a consultant to, MicroGenDX at the time of submission.

## ABBREVIATIONS

CLR: centered-log ratio
EV1: eigenvector 1
EV2: eigenvector 2
GTEx: Genotype-Tissue Expression
mbGWAS: microbiome genome-wide association study
mbTWAS: microbiome transcriptome-wide association study
OTU: operational taxonomic unit
SNPs: single nucleotide polymorphisms

